# Circulating tumor fraction analyses with ultra-low pass whole genome sequencing predict response to chemoradiation and recurrence in stage IV small cell carcinoma of the cervix: a longitudinal case study

**DOI:** 10.1101/2020.11.18.20233502

**Authors:** Ata Abbas, Morgan Gruner, Jennifer Karohl, Peter G. Rose, Amy Joehlin-Price, Hussein Al-Sudani, Daniel Stover, Haider Mahdi

**Affiliations:** Division of Hematology and Oncology, Department of Medicine, Case Western Reserve University, Cleveland, OH 44106; Division of Gynecologic Oncology; Obstetrics, Gynecology and Women’s Health Institute, Cleveland Clinic, 9500 Euclid Avenue, Cleveland, OH 44195; Department of Anatomic Pathology, Pathology and Laboratory Medicine Institute, Cleveland Clinic, 9500 Euclid Avenue, Cleveland, OH 44195; Department of Internal Medicine, Albert Einstein Healthcare Network, Philadelphia, PA 19403; Division of Medical Oncology, The Stefanie Spielman Comprehensive Breast Center, The James Cancer Hospital and Solove Research Institute at The Ohio State University Wexner Medical Center, Columbus, OH; Translational Hematology Oncology Research Department, Taussig Cancer Institute, Cleveland Clinic, 9500 Euclid Avenue, Cleveland, OH 44195; Genomic Medicine Institute, Lerner Research Institute, Cleveland Clinic, 9500 Euclid Avenue, Cleveland, OH 44195

**Keywords:** Advanced Stage Small Cell Carcinoma of Cervix, circulating tumor DNA (ctDNA), biomarker, ultra-low spass whole genome sequencing

## Abstract

Neuroendocrine carcinoma of the cervix is a rare and aggressive form of cervical cancer that presents with frequent metastasis at diagnosis and high recurrence rates. Primary treatment is multimodal, which often includes chemotherapy with or without radiation therapy. There is no data available to guide treatment for recurrence, and second-line therapies are extrapolated from small cell lung carcinoma data. Close monitoring of these patients for recurrence is paramount. Evaluation of circulating tumor DNA (ctDNA) in the peripheral blood is an attractive approach due to its noninvasive nature. Ultra-low pass whole genome sequencing (ULP-WGS) can assess tumor burden, response to therapy, and predict recurrence; however, data are lacking regarding the role of ULP-WGS in small cell carcinoma of the cervix. This study demonstrates a patient whose response to chemotherapy and cancer recurrence was accurately monitored by ctDNA using ULP-WGS testing and confirmed with radiologic imaging findings.

## INTRODUCTION

Neuroendocrine carcinoma of the cervix is a rare form of cervical cancer and accounts for only 2% of all invasive cervical cancers ^1,2^. The majority are small cell carcinomas, which tend to behave aggressively, are characterized by a high mitotic rate, extensive necrosis, high propensity for lymphovascular space invasion (LVSI), and frequently associated with HPV 18 ^3,4^. Neuroendocrine carcinomas of the cervix behave with an aggressive nature; therefore, they require a multimodality treatment approach, which often includes platinum/etoposide-based chemotherapy with or without radiation ^5^. Gardner et al. published the SGO clinical document that provided treatment guidelines highlighting radical excision and chemotherapy for early-stage disease ^6^. It tends to affect young women with a median age of 37 years. The prognosis is poor, usually a short clinical course with a median survival of less than 24 months ^7,8^. In patients with recurrent disease, data to guide treatment decisions are absent for this rare cancer subtype. Second-line drugs are mostly extrapolated from small cell lung cancer, including chemotherapy like topotecan, paclitaxel, or VAC combination chemotherapy. However, these regimens are toxic with limited activity. Recently, the role of immunotherapy with immune checkpoint inhibition has been explored in small cell lung cancer and other high-grade neuroendocrine carcinomas ^9,10^. Even though the benefit is restricted to small subgroups, the data is limited in NEC of the cervix or other gynecologic origin. Recently, Frumovitz et al. reported low activity of pembrolizumab in cases series of 7 patients with recurrent small cell carcinoma of the cervix and other lower female genital tract ^11^.

Evaluation of circulating tumor DNA (ctDNA) in the peripheral blood of patients with solid tumors represents an attractive approach given its non-invasive nature and the potential to do a serial evaluation at the time of diagnosis, during therapy, and follow up ^12-14^. However, the extent of the shedding of tumor DNA into peripheral blood depends on many factors, including the type of cancer, the extend of disease, visceral metastasis, etc. The impact of ctDNA on the management of solid tumors has been highlighted in several aspects, including somatic genomic characterization, therapy selection, and tumor fraction as well as molecular residual disease to predict tumor burden, prognosis, response and resistance to therapy, and disease recurrence ^14-17^.

Ultra-low pass whole genome sequencing (ULP-WGS) represents an attractive approach to assess tumor burden and therapeutic response/resistance, given that it is affordable, cost-effective, and does assess the whole genome with low coverage (0.1 x coverage) ^17,18^. The data on the role of ctDNA in general and specifically ULP-WGS in predicting prognosis, response to chemoradiation, and recurrence in small cell carcinoma of the cervix are lacking. ULP-WGS could be a potentially promising approach given the aggressive nature of this cancer and the tendency for metastatic spread at the time of diagnosis and recurrences. Further, these patients tend to have a scant amount of tissue that limits extensive genomic characterization because most of them are treated with chemotherapy or chemoradiation rather than surgical resection.

In this study, we sought to investigate the tumor fraction and copy number alteration in ctDNA using ULP-WGS in peripheral blood of a patient diagnosed with stage IV small cell carcinoma of the cervix who underwent chemotherapy with cisplatin and etoposide concurrent with pelvic radiation and achieved completed clinical response followed by recurrent disease in the liver, adrenal gland, and brain within six months. Here, we show that tumor fraction assessment at the time of diagnosis and during treatment predicted response to chemoradiation therapy and recurrent disease.

## RESULTS

### Clinical case course

In her early thirties, a woman with a noncontributory past medical history presented to medical care with the complaint of brown vaginal discharge and an abnormal sensation in her vagina. A 2.5-3 cm pedunculated mass was noted on examination with cervical biopsy pathology positive for invasive high-grade neuroendocrine carcinoma, most consistent with small cell type, associated with endocervical adenocarcinoma in situ. Immunostaining for p63, p40, synaptophysin, and chromogranin was performed to assist with cell typing. The carcinoma cells express neuroendocrine markers synaptophysin and chromogranin (Figure 1A-B). P63 stains were weakly positive, and the p40 stain was negative (data not shown). She then underwent MRI and PET/CT imaging, which was positive for uptake in the cervix region. Metastases in the liver and right adrenal gland were also noted (Figure 1C), leading to the final diagnosis of Stage IV invasive high-grade neuroendocrine carcinoma of the cervix, small cell type. The patient began therapy with Cisplatin 75 D1 and Etoposide D1,2,3 –Q21 days. She was transitioned to Carboplatin AUC 5 after C2 due to severe tinnitus. After three cycles of the above chemotherapy, a CT scan was performed, which demonstrated decreased hepatic metastasis and complete resolution of a left adrenal metastatic lesion. The patient received four additional cycles of the aforementioned Carboplatin and Etoposide and then underwent a repeat PET/CT scan, which demonstrated complete interval resolution of the previously seen hypermetabolic cervical mass, hepatic mass, and adrenal mass (Figure 2A). The patient then underwent two additional cycles (consolidation cycles) of Carboplatin and Etoposide after imaging confirmation of disease resolution. Simultaneously, during the consolidation chemotherapy cycles, the patient underwent EBRT to the pelvis with a total dose of 4500 cGy in 25 fractions and HDR brachytherapy to cervix and parametria with a total dose of 1400 cGy in 2 fractions. The patient completed therapy and was with no evidence of disease for five months. One year after her initial diagnosis and five months after completing therapy, she presented with new-onset dizziness. She underwent imaging, which demonstrated an interval development of hepatic and right adrenal metastatic disease (Figure 2B, right) as well as a heterogeneously enhancing 2.9 cm likely intra-axial mass involving the right temporal lobe (Figure 2B, left). She received a gamma knife to two brain targets – the right temporal lobe and left insular regions. She was then started on Ipilimumab plus Nivolumab immunotherapy. Newly detected spinal nerve root metastases and leptomeningeal disease were discovered, for which she underwent 20 Gy in 5 fractions of palliative RT to spine. Shortly thereafter, the patient was admitted to the hospital in the setting of altered mental status and noted to have disease progression. The family selected hospice care, and the patient expired 15 months after the initial diagnosis.

**Figure 1.**
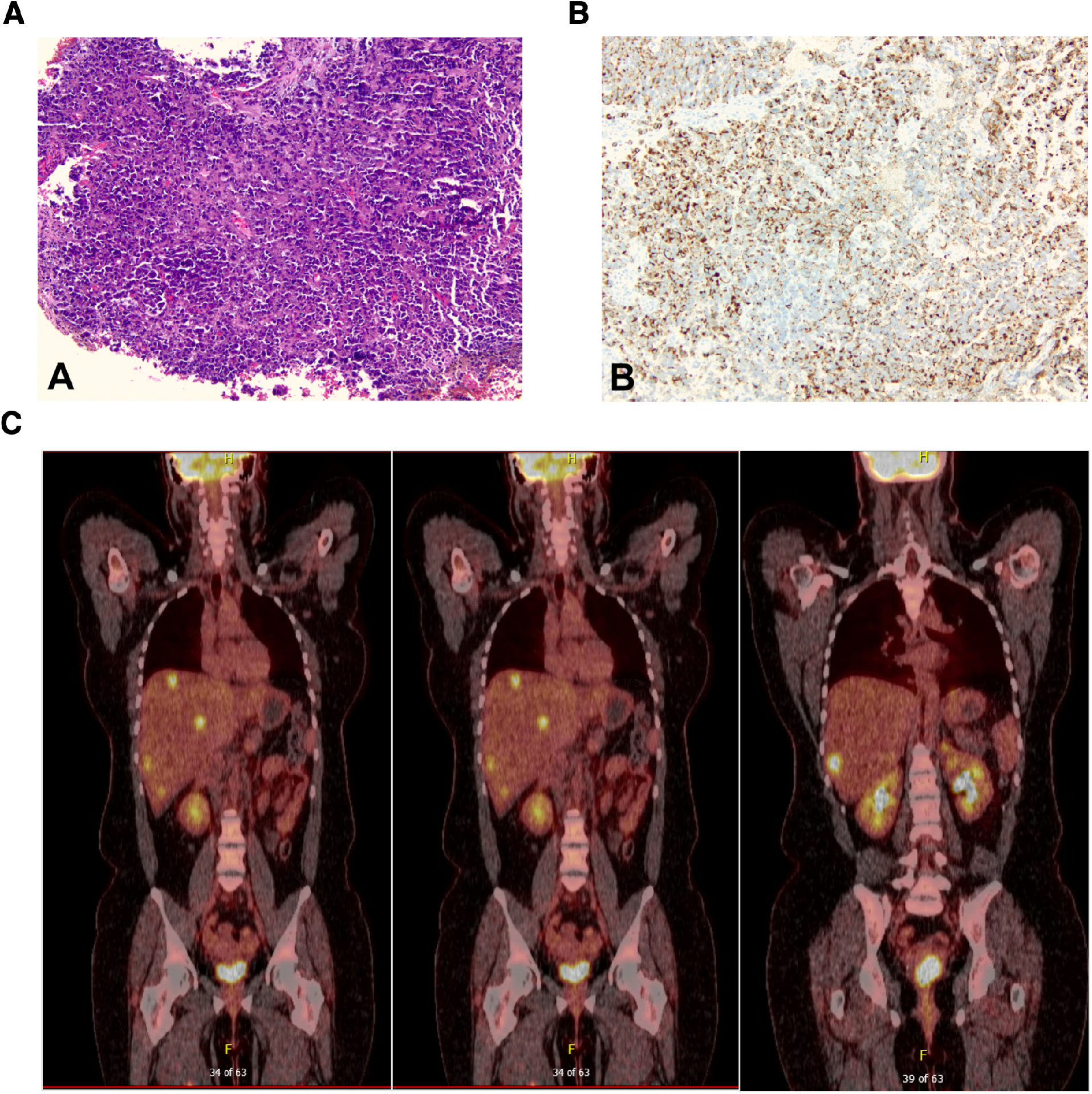
**(A)** The initial cervical biopsy (100x magnification) demonstrated extensive mitotic activity, high-grade nuclei with hyperchromasia and nuclear molding, scant cytoplasm, and ill-defined cell borders, all characteristic of small cell carcinoma. **(B)** The carcinoma was diffusely positive for chromogranin (100x magnification). **(C)** Positron Emission Tomography/Computed tomography (PET/CT) scan at the time of diagnosis that showed evidence of cervical disease, multiple hepatic metastases, and right adrenal gland metastasis.

**Figure 2.**
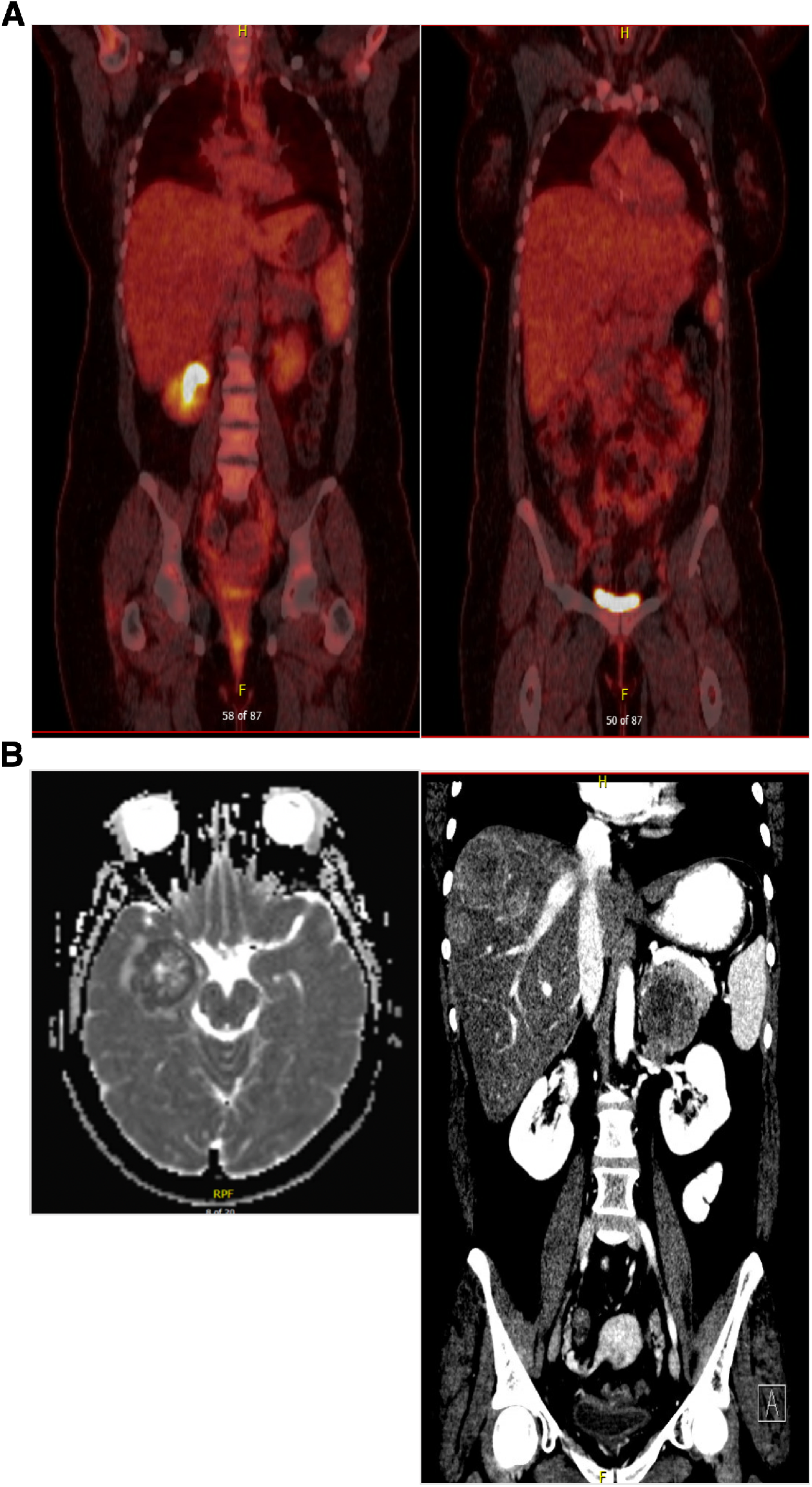
**(A)** Positron Emission Tomography/Computed tomography after completing chemoradiation therapy showed no evidence of disease. **(B)** Computed tomography and Magnetic resonance imaging of the brain and computed tomography of the abdomen at the time of diagnosis of recurrent disease confirming brain metastasis and multiple hepatic metastatic disease.

### Correlation of tumor fraction (TFx) by ULP-WGS with clinical outcome

To understand the role of ctDNA using TFx as a non-invasive biomarker, first, we sought to investigate the relationship of TFx with disease burden at the time of diagnosis and during chemoradiation. We focused on the time of diagnosis, during treatment, and at the completion of treatment. At the time of diagnosis, her tumor TFx was 26%, with wide variability in somatic copy number alteration (sCNA). After completion of chemotherapy, her TFx dropped to 5%, which coincided with the finding of complete response by her PET/CT scan. Then after completion of pelvic radiation a month after, her TFx was 2.5%. Then five months later, she had another evaluation, and her TFx was 36%. This was consistent with the diagnosis of recurrent disease that was evident by imaging confirming recurrent brain, multiple hepatic, and right adrenal metastasis (Figure 3A and 3B).

**Figure 3.**
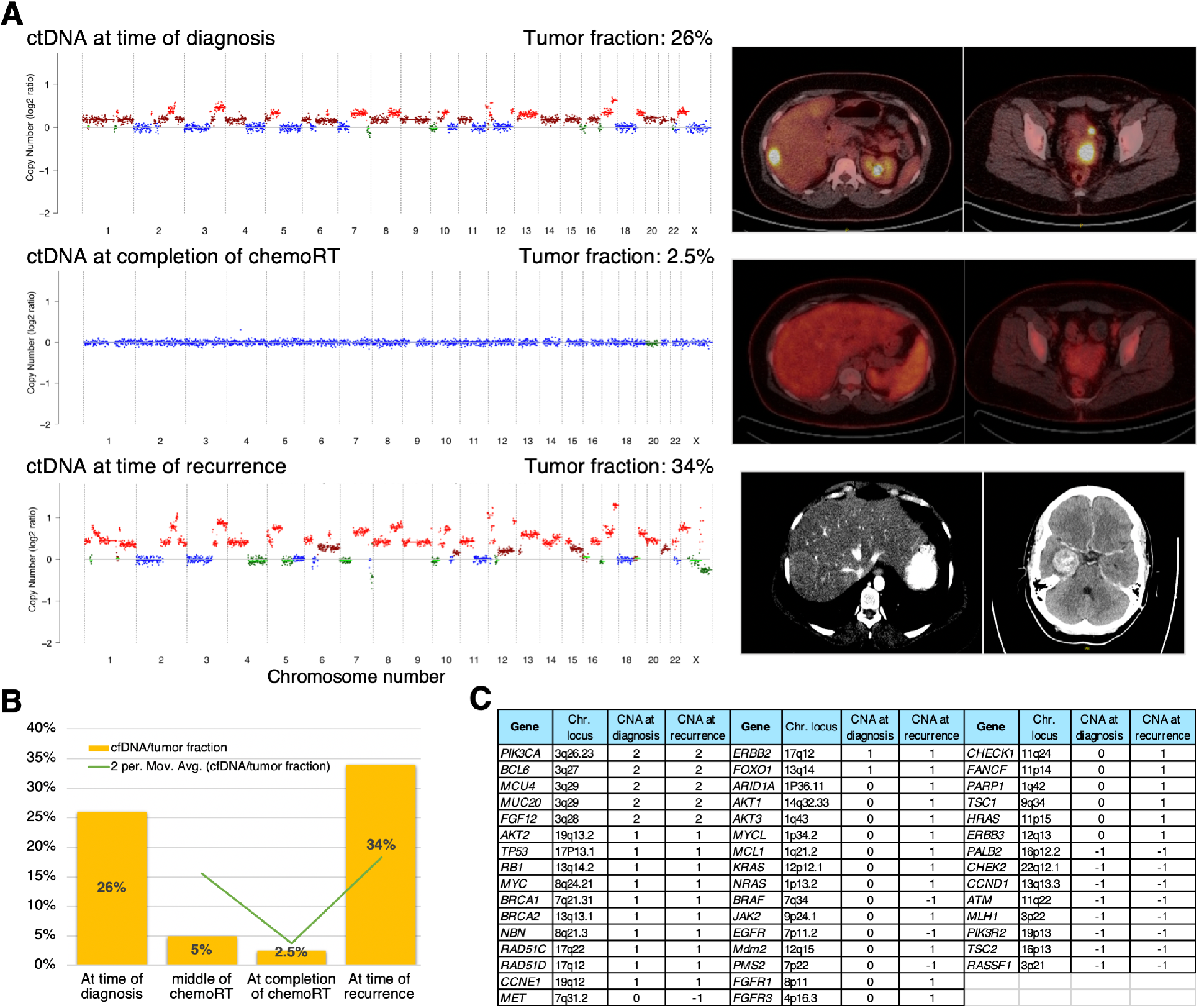
**(A)** Copy number plots of three representative ctDNA (left panels) with copy number (log2 ratio) indicated on the y-axis and chromosome on the x-axis. Positron emission tomography/Computed tomography at the three representative time points corresponding to the time of collection of ctDNA samples (right panels). **(B)** diagrammatic representation of the serial trend of ctDNA/tumor fraction at diagnosis, during treatment, and at time of recurrence. **(C)** A descriptive comparison of selected gene-level copy number data between time of diagnosis and time of recurrence. 2 / -1 / 0 / 1 / 2 refer for homozygous deletion, heterozygous deletion, copy neutral, low gain, high amplification, respectively.

### Comparison of somatic copy number alteration (sCNA) and gene-levels copy number alteration with ULP-WGS between primary disease at initial diagnosis and recurrent disease

Then we sought to investigate sCNA at these above time points and to see if there is a difference between the primary and recurrent disease and if there are interval changes. sCNA at the time of recurrence was comparable to that at the time of initial diagnosis but with higher amplitude, especially for amplification (Figure 3A). Similarly, genes-level copy number data were comparable too with evidence of additional gains at the time of recurrence as evident in selected genes that correspond to certain pathways like PI3K/AKT, RAS/RAF, DNA-damage response, MYC/MCL, TP53 and RB1 as well as other pathways (Figure 3C).

## DISCUSSION

Here, we present a case study with longitudinal monitoring of tumor fraction in ctDNA using ULP-WGS in peripheral blood of a patient diagnosed with stage IVB small cell carcinoma of the cervix. We demonstrate that tumor fraction correlates positively with radiologic disease burden and response to chemoradiation therapy. Our data show evidence of tumor shedding into peripheral blood at the time of diagnosis in metastatic small cell carcinoma of the cervix. At the time of complete metabolic response by PET/CT scan, the tumor fraction declined from 26% to 2.5%, which confirm the potential in predicting response to therapy. Furthermore, tumor fraction spiked to 36% at the time of recurrent disease, which correlated positively with the radiologic evidence of recurrent metastatic disease. This highlights the potential role of monitoring tumor fractions with ULP-WGS technology to monitor tumor burden, response to therapy, and disease recurrence in high-grade neuroendocrine carcinoma of the cervix.

This is the first report to the best of our knowledge that highlights the role of serial assessment of tumor fraction with ULP-WGS in this cancer. Small cell carcinoma of the cervix is rare, very aggressive, and has a poor prognosis ^3,4,7,8^. ctDNA may play a role in monitoring disease burden, assessing response to therapy, and predicting disease recurrence during surveillance ^12-17^. It is likely a more acceptable approach to these patients, especially during surveillance, due to the overall ease and safer blood draw. Further, these cancers tend to be metastatic at the time of diagnosis and less likely to be treated with surgical resection. Given that, getting adequate tissue is usually a problem, and often, there is scant tissue from biopsy to obtain a diagnosis. Therefore, ctDNA can be used to genomically characterize these cancers at diagnosis and monitor recurrence and disease progression.

The potential explanation for high tumor fraction at the time of diagnosis and recurrence is the presence of distant visceral metastasis, especially to the liver, high disease burden, and aggressive biology of this cancer. Prior studies reported that liver metastasis is associated with more shedding of ctDNA than other metastatic sites, which could be explained by the anatomy and blood flow supporting the metastatic disease in the visceral sites ^17,19^. The potential utility is to monitor patients during surveillance for evidence of visceral metastasis, especially when other diagnostic modalities are negative like imaging and other biomarkers or those with rising biomarkers with negative imaging, which represent a real challenge in some cancers.

Another critical question is whether this patient had a complete molecular response or not at the completion of therapy. Although she had a complete metabolic response with full resolution of her disease, it is not clear if there was still residual molecular disease given that the tumor fraction significantly dropped to 2.5%. It is not clear whether there is an optimal cutoff that can be used to make a judgment on the role of therapy duration, treatment intensification, or changing treatment regimens. These need to be investigated further in future prospective studies.

There is no reliable serologic biomarker to monitor response to therapy or recurrence during the surveillance of patients with small cell carcinoma of the cervix. Therefore, Longitudinal monitoring of ctDNA represents an attractive approach in managing these patients, especially if combined with radiology imaging. Interestingly, the pattern of copy number alteration at the time of recurrence was comparable to that of initial diagnosis but with higher amplitude, and genes-level data were consistent with that. These data support that this aggressive cancer at time of recurrence have similar molecular characteristics compared to primary disease but with higher amplitude of amplification at least at the copy number level. These data are interesting but should be interpreted cautiously and need to be validated in the future in a prospective cohort.

In conclusion, this case study demonstrates the role of serial assessment of tumor fraction and ctDNA as a potentially promising non-invasive biomarker in patients with advanced-stage small cell carcinoma of the cervix. Future studies are warranted to assess the role of ctDNA as a prognostic and predictive biomarker in this highly aggressive cancer, particularly in correlation with the extent of disease, tumor burden, and site of metastasis. Further, it will be important to explore the additional benefit if ctDNA combined with radiology imaging during follow up of response to therapy and routine surveillance.

## METHODS

### Clinical case data extraction

The clinicopathologic, treatment administration as well as treatment duration and response evaluation data were collected retrospectively from the patient’s chart. Patients have consented for blood collection prospectively, and it was by the institutional review board.

### Clinical specimens, circulating tumor DNA extraction and quantification

Plasma samples were prospectively collected as part of an ongoing study approved by the Cleveland Clinic Institutional Review Board. Blood was collected in 10 ml of Cell-Free DNA BCT (Streck, Omaha, NE). Blood was processed to collect plasma and buffy coat through standard density gradient centrifuge protocol. In brief, blood samples are centrifuged at 1500 rpm for 6 minutes, the plasma is isolated and stored in 1.5 ml microfuge tubes and frozen in -80°C till further processing. Further, DNA was extracted using the standard protocol. In brief, RBC lysis buffer was used for 5-10 minutes then spun at 450 RCF for 5 minutes, the supernatant was removed, and 500 ml of PBS was added and spun for 5 minutes then lysis solution was added to lyse overnight, and then samples were stored in -80°C till future use. Subsequently, for the purpose of ctDNA analysis, frozen aliquots of the plasma samples were thawed at room temperature and subjected to a second high-speed spin after thawing. Cell-free DNA extraction and quantification were performed as previously described ^12^.

### Ultra-Low-Pass Whole-Genome Sequencing (ULP-WGS)

Library construction of ctDNA was performed using the Kapa HyperPrep kit with custom adapters (IDT, Coralville, IA). Three to 20 ng of ctDNA input (median, 5 ng), or approximately 1,000 to 7,000 haploid genome equivalents, was used for ultra-low-pass whole-genome sequencing. Constructed sequencing libraries were pooled and sequenced using 100-bp paired-end runs on a HiSeq2500 (Illumina, San Diego, CA) to get an average genome-wide fold coverage of 0.13. Segment copy number and tumor fraction (TFx) were derived via ichorCNA ^12^. Samples were excluded if the median absolute deviation of copy ratios (2^log2 ratio^) between adjacent bins, genome-wide, was > 0.20, suggesting poor-quality sequence data.

### Gene-Level Copy Number Analyses

GISTIC2.0 output ^20,21^ was used for all gene-level copy number analyses. Segmented data files derived from ichorCNA were purity and ploidy corrected, then input into GISTIC2.0 with amplification/ deletion threshold log_2_ratio > 0.3, confidence level 0.99, and Q-value threshold 0.05. Genes were defined as gain (GISTIC value 1; corresponds to three copies) or amplification (GISTIC value 2; corresponds to four or more copies) versus diploid (GISTIC value 0).

## Data Availability

The data are available from the corresponding author on reasonable request.

## DATA AVAILABILITY

The Ultra-low pass whole genomic sequencing dataset generated during the current study (for all the time points) are not publicly available as these are patient samples with potentially identifiable germline SNPs and there is no patient consent for depositing this sequencing data in a public repository. However, the data are available from the corresponding author on reasonable request.

